# Early intravenous Beta-Blockade with esmolol in adults with isolated severe Traumatic Brain Injury (EBB-TBI): protocol for a phase 2a intervention design study

**DOI:** 10.1101/2023.02.27.23286185

**Authors:** Matt Thomas, Kati Hayes, Paul White, Aravind Ramesh, Lucy Culliford, Gareth Ackland, Anthony Pickering

**Affiliations:** North Bristol NHS Trust; School of Data Science and Mathematics, University of the West of England, Bristol; Faculty of Health Sciences, University of Bristol; Bristol Trials Centre, Bristol Medical School (PHS), University of Bristol; Translational Medicine and Therapeutics, William Harvey Research Institute, Queen Mary University of London; School of Physiology, Pharmacology and Neuroscience, University of Bristol

**Author notes:** Corresponding author Dr Matt Thomas, Intensive Care Unit, Southmead Hospital, Bristol, BS10 5NB.

**Keywords:** Brain injuries, traumatic, adrenergic beta-antagonists, intensive care units, models, statistical

## Abstract

Traumatic brain injury is a leading cause of death and disability worldwide. Interventions that mitigate secondary brain injury have the potential to improve outcomes for patients and reduce the impact on communities and society. Increased circulating catecholamines are associated with worse outcomes and there is supportive animal data and indications in human studies of benefit from beta-blockade after severe traumatic brain injury. Here we present the protocol for a dose-finding study using esmolol in adults commenced within 24 hours of severe traumatic brain injury. Esmolol has practical advantages and theoretical benefits as a neuroprotective agent in this setting, but these must be balanced against the known risk of secondary injury from hypotension. The aim of this study is to determine a dose schedule for esmolol, using the continual reassessment method, that combines a clinically significant reduction in heart rate as a surrogate for catecholamine drive with maintenance of cerebral perfusion pressure. The maximum tolerated dosing schedule for esmolol can then be tested for patient benefit in subsequent randomised controlled trials.

## Introduction

Traumatic brain injury (TBI) is a global public health emergency. It is the leading cause of death in young adults and a major cause of death and disability in all ages worldwide. The impact is felt by patients, families and communities. In the United Kingdom (UK) the annual cost is estimated at £15 billion with more than 68,000 years of life lost [1,2].

Neuroprotection is the preservation, salvage or recovery of central nervous system function after acute insult. Better neuroprotective strategies would reduce death and disability after TBI. The current management strategies are conservative, relying on provision of physiological stability and timely management of complications (e.g., seizures, intracranial hypertension). There is no proven therapy that provides additional benefit [3].

There are plausible reasons to consider beta-blockers as potential neuroprotective drugs after traumatic brain injury. Plasma catecholamine levels correlate with inflammation, endothelial injury and poor outcome after TBI [4,5,6]. In animals, administering beta-blockers after TBI reduced cerebral oedema and hypoxia (mice), increased perfusion (mice), protected cerebral autoregulation (pigs) and was associated with better recovery (mice) [7,8,9].

In patients a number of meta-analyses show that there is potential benefit for beta-blockade in traumatic brain injury to reduce mortality and improve functional outcomes [10,11,12,13,14]. However, the studies are heterogenous and largely observational. A few small randomised trials are supportive but have limited external validity and/or have not been subject to peer review [15,16,17,18]. This means there is still uncertainty about the overall effectiveness of beta-blockade as well as the specifics of patient selection, drug, dose, route and physiological goal.

Repurposing an established beta-blocker for neuroprotection could significantly improve individuals’ health outcomes, reduce the impact on families and communities, and save resources for health systems and societies at a low cost, potentially representing excellent value for money. There is though a risk with the use of anti-hypertensive drugs early after severe traumatic brain injury; compromising blood pressure maintenance in the TBI patient group, for whom hypotension has been called “the single most important secondary insult”, could lead to worse outcomes [19].

With this risk-benefit balance in mind we designed the Early Beta Blockade in adults after severe Traumatic Brain Injury (EBB-TBI) programme, of which this is the first of three planned studies. The overarching hypothesis of the EBB-TBI programme of research is that beta-1 adrenoceptor blockade after severe traumatic brain injury in adults reduces morbidity and mortality by reducing secondary brain injury driven by the hyperadrenergic state. Here we describe the protocol for the first study. We use methodology common to early phase drug studies to test escalating starting doses for esmolol infusion in small cohorts of patients with an end point of heart rate control without compromise of cerebral perfusion pressure. The aim is to optimise the intervention with esmolol so it can then be tested in subsequent trials of efficacy and effectiveness.

### Rationale for choice of esmolol and dosing regime

There are several characteristics of esmolol that are well suited to use for beta-blockade in the early phase of critical illness. First and foremost, the rapid onset (time to steady state of action of 5 minutes), facilitates titration to the desired clinical endpoints. This, coupled with the short elimination half-life of 9 minutes means rapid offset of effect if reduced or stopped for adverse events. The drug has few interactions with other medications beyond the general class effects of beta-1 selective-blockers. It is metabolised to inactive compounds and this is independent of organ function. The intravenous route of administration bypasses the gut and guarantees drug delivery in a population where gastric emptying is frequently delayed. Finally, it is inexpensive, in common use in intensive care units and preparation and administration are straightforward (e.g., it is stable at room temperature for up to 24 hours, protection from light is not required and it can be administered via peripheral venous access) [20,21].

While the primary reason for choosing esmolol is practical there are also theoretical arguments. A beta-1 selective blocker (metoprolol) is used in the Lund approach to the management of TBI as it does not cause cerebral vasodilatation or alter cerebral blood flow after severe traumatic brain injury [22,23]. Esmolol itself has been shown to have no adverse effect on cerebral blood flow in volunteers or in patients receiving electroconvulsive therapy [24,25]. In the setting of anaesthesia, with the same drugs commonly used for sedation after TBI, esmolol provides additional cortical suppression that could contribute to neuroprotection [26]. In animals esmolol is neuroprotective after brain or spinal cord ischaemia [27,28]. Finally there is evidence that esmolol reduces the inflammatory response after surgical trauma [29].

The initial dose selected for testing in the first cohorts in the EBB-TBI study (esmolol infusion started at 5 micrograms per kilogram per minute) is based on a regimen shown to be tolerated by a population with severe septic shock who are, like many patients after severe traumatic brain injury, mechanically ventilated and receiving vasopressors [30]. The esmolol was then titrated against heart rate using dose increments of 2.5 micrograms per kilogram per minute (mcg.kg.min^-1^) every 30 minutes. The infusion starting rate for subsequent cohorts was determined by using the “so-called modified Fibonacci sequence” (commonly used in oncology dose-finding studies [31,32]) using the initial EBB-TBI cohort dose as the start of the sequence.

Doses as low as 15 micrograms per kilogram per minute (mcg.kg.min^-1^) are anti-inflammatory after surgical trauma in humans [29]. The EC50 for reduction in heart rate during exercise in humans is 113 mcg.kg.min^-1^[33]. Side effects, mainly hypotension, are more common with doses exceeding 150 mcg.kg.min^-1^[20,33]. In rats a dose of 20 mcg.kg.min^-1^ is protective against cerebral ischaemia, and 16 mcg.kg.min^-1^ is anti-inflammatory in sepsis [34,35]. While the minimum dose required for neuroprotection in humans is not known this supports the concept that a dose of esmolol resulting in heart rate reduction is also potentially effective in reducing brain injury, and that neuroprotection might be possible while avoiding serious side effects.

Rather than a fixed dose, the infusion will be titrated to heart rate, a convenient clinical biomarker for sympathetic nervous system activity and catecholamine drive, allowing for personalisation of dose. We will aim for a 15% reduction from pre-enrolment baseline. Titration of medication to achieve physiological goals is routine in intensive care practice. Use of heart rate avoids any conflict with accepted physiological goals like blood pressure set in clinical guidelines (such as those from the Brain Trauma Foundation [19]).

In healthy volunteers heart rate reduction of 15% induced by esmolol did not alter cerebral blood flow [24]. After ST elevation myocardial infarction a 14% reduction in heart rate with esmolol did not increase the incidence of cardiogenic shock or atrio -ventricular block while limiting the peak cardiac troponin T release [36]. A 20% reduction in heart rate in septic mechanically ventilated patients did not alter oxygen utilisation or hepatic or leg blood flow [37]. Evidence relating to the ideal target heart rate in severe traumatic brain injury is conflicting [38,39].

We aim to administer esmolol as soon as practicable after injury, to achieve early blockade of the hyperadrenergic surge accompanying the head trauma and interruption of self-perpetuating pathophysiological cascades before secondary injury is established [40]. Observational data shows catecholamine related pathophysiology at the time of hospital admission for TBI [5]. The duration of the infusion is limited to four days from enrolment. This translates into an intervention period that spans the time of initial development of cerebral oedema after injury. This regimen is therefore based on a combination of clinical judgement of the time of greatest sympathetic activation with knowledge of the typical course of intracranial hypertension [41,42]. It allows the primary research question for this study to be answered at the time of greatest potential haemodynamic instability without unnecessarily prolonged exposure to a drug that may not ultimately provide benefit. After this period, the continued use of esmolol or any other selective or non-selective beta-blocker is at the discretion of the treating clinician. To the best of our knowledge there is no literature examining the optimal timing of administration of beta-blockers after traumatic, ischaemic or septic insult.

### Rationale for use of continual reassessment method

Dose-finding studies are used in early phase research to estimate the dose-toxicity profile of drugs and to select the right dose for subsequent trials. Rule-based study designs rely on pre-defined rules to determine future dosage decisions based on observed toxicity at current doses. In contrast, model-based designs use statistical models to guide these decisions based on a target level of toxicity that combines judgments of the potential benefit of drug administration and severity of harm arising from toxicity.

The continual reassessment method (CRM) is a parametric model-based study design. Several advantages are reported for model-based designs like the CRM over traditional rule-based designs, including flexibility and increased precision. This aids an efficient study design that will minimise the number of patients required to determine the maximum tolerated dose and maximises safety by exposing the fewest patients possible to either under- or over-treatment and by rapid dose titration [43,44,45,46].

Additional safety measures are possible with the CRM without compromise of study performance [47]. Given the lack of data on early esmolol dosing in traumatic brain injury, the uncertain magnitude of any benefit and the potential for severe harm with toxicity (i.e. hypotension) this is important for this study. A run-in cohort of three patients, increased number of cohorts, and avoidance of skipping untried doses have been incorporated in this protocol.

### Study outline

This is a prospective dose-finding study of esmolol aiming to attenuate the sympathetic surge associated with severe traumatic brain injury in adults. The setting is the 48-bed mixed Intensive Care Unit of Southmead Hospital, Bristol, a 996-bed teaching hospital in the South West of England and a Major Trauma Centre serving an adult population of approximately 2.3 million.

The primary objective is to define a treatment dose escalation schedule for esmolol for use in adults early (<24 hours) after severe traumatic brain injury that balances the potential benefit of early exposure to beta-blockade with the ability to maintain adequate cerebral perfusion pressure. A reduction of heart rate of at least 15% from pre-infusion baseline will be used as an indicator of clinically significant beta-blockade.

A group sequential adaptive model-based design (the continual reassessment method) will be used to determine the maximum tolerated dose schedule for esmolol, defined as the highest dosing regimen associated with an acceptable level of toxicity. For the purpose of this study this is taken to be a probability of dose-limiting toxicity of 10%. Dose-limiting toxicity is defined as failure to maintain cerebral perfusion pressure above the minimum recommended by Brain Trauma Foundation guidelines (60 mmHg) despite standard interventions and the protocolised de-escalation of the esmolol infusion such that withdrawal of esmolol is required; or the occurrence of a serious adverse event mandating withdrawal of the esmolol infusion.

Secondary objectives are to identify the effects of esmolol on organ function and to record clinical outcomes including mortality and function at 6 months. Exploratory objectives are to monitor the effects on biomarkers of sympathetic nervous system activity and to gather data to inform the design of a randomised controlled trial to establish feasibility and efficacy.

### Study flow chart

Flow through the study is shown in Figure 1.

**Figure 1:**
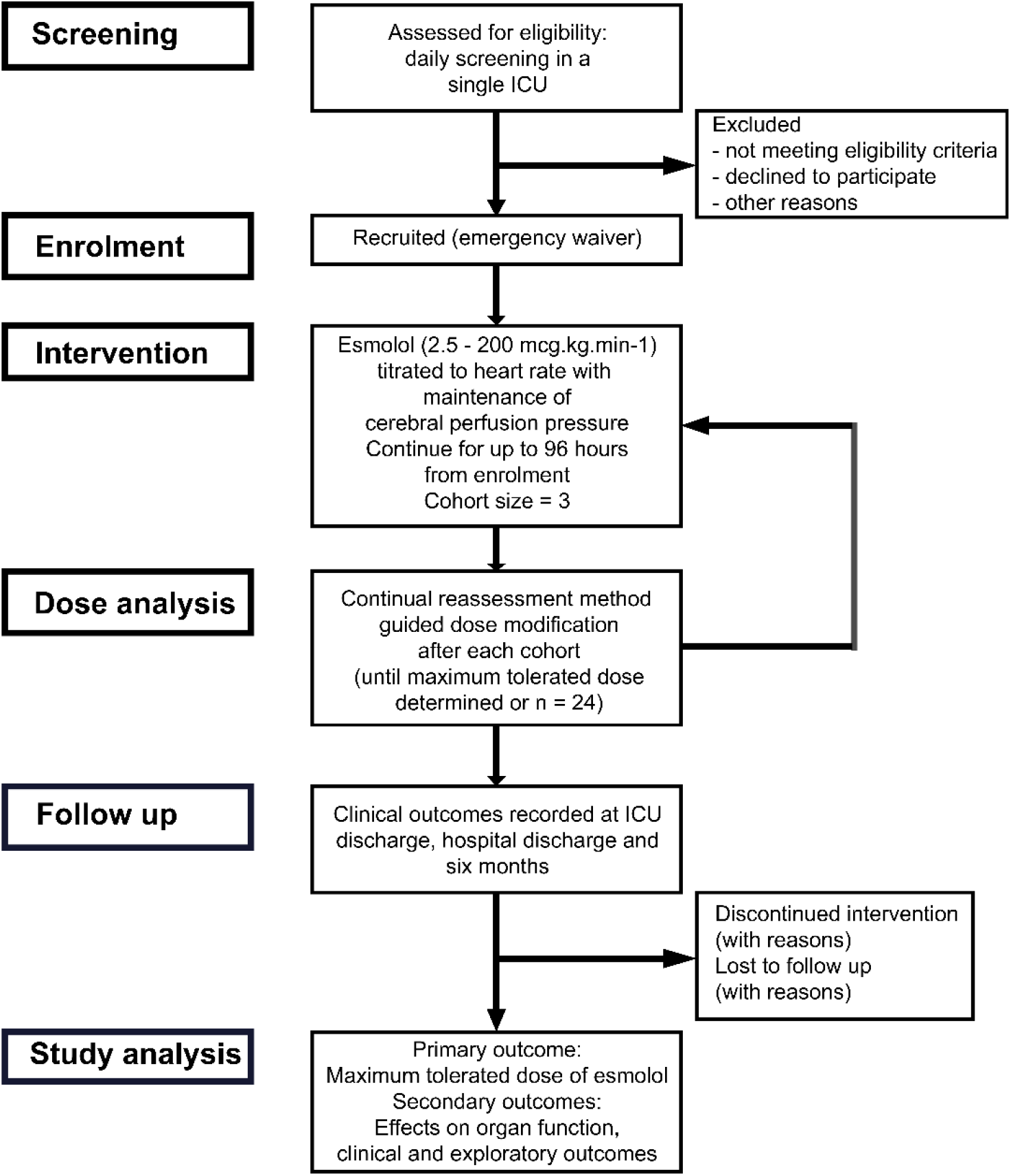
Flow of patients through the study. ICU – Intensive Care Unit; mcg.kg.min^-1^ – micrograms per kilogram per minute.

### Eligibility

Participants must meet all inclusion criteria and none of the exclusion criteria (shown in Table 1) and start the esmolol infusion within 2 hours of confirmation of eligibility. The baseline heart rate must be > 60 beats per minute for more than 15 minutes for the infusion to start.

**Table 1:** eligibility criteria. AV – atrio-ventricular; mmHg – millimetres of mercury; NYHA – New York Heart Association; PA – pulmonary artery.

| Inclusion criteria (all to be met) | Exclusion criteria (none to be met) |
| --- | --- |
| 1. Aged 18 years or over<br>2. Severe traumatic brain injury (Glasgow Coma Score of 8 or less after resuscitation of prior to intubation)<br>3. Within 24 hours of injury<br>4. Intracranial pressure monitoring in situ | 1. Life of limb threatening extra-cranial injury (in the opinion of the treating intensivist)<br>2. Perceived devastating brain injury admitted for the purposes of prognostication or organ donation<br>3. Participation in another clinical trial of investigational medicinal product within preceding 30 days<br>4. Pregnancy<br>5. Breast feeding<br>6. Hypersensitivity to beta-blockers<br>7. Cardiogenic shock<br>8. Decompensated heart failure (NYHA class 4)<br>9. Untreated sick sinus syndrome or AV nodal conduction disorders including second or third degree heart block)<br>10. Untreated phaeochromocytoma<br>11. Acute severe bronchospasm<br>12. Severe pulmonary hypertension (mean PA pressure >55mmHg)<br>13. Prinzmetal's angina<br>14. Severe metabolic acidosis (pH < 7.1)<br>15. Use of verapamil within the preceding 48 hours |

These criteria exclude those at greatest risk of harm from beta-blockade, target early enrolment for greatest potential benefit and remain broad to capture a representative patient population.

### Consent

Potentially eligible participants, as a consequence of their severe traumatic brain injury, will lack the capacity to provide consent for this study. The need for early intervention to maximise potential benefit, and the uncertainty of time and extent of recovery, means it is not practicable to wait for capacity to return. The time critical nature of the intervention and the potential for significant additional distress in the emergency situation precludes seeking prior Personal Legal Representative opinion. As such an emergency waiver of consent model will be used, with informed consent sought once patients regain capacity, as laid out in UK legislation (The Medicines for Human Use (Clinical Trials) Amendment (No.2) Regulations 2006).

Although any participant (or their Legal Representative) may withdraw their consent at any time, given the adaptive nature of the study with frequent analysis of patient data and the dependence of later study drug dosing on prior patient response it is not feasible to withdraw all data unreservedly. Participant data that has been used in study drug dose calculation will not be withdrawn.

### Study intervention

Open-label esmolol is given as a continuous intravenous infusion at a dose of 2.5 – 200 micrograms per kilogram per minute (mcg.kg.min^-1^) titrated using a predefined dose escalation schedule to achieve a heart rate reduction of ≥15% from baseline with cerebral perfusion pressure maintained above 60mmHg. The target heart rate will be set as the rolling mean over the preceding 5 minutes to avoid overshoots. Baseline is defined as the mean heart rate in the 4 hours preceding confirmation of eligibility. The minimum permitted target heart rate will be 60 beats per minute (bpm) and maximum will be 100 beats per minute. Actual body weight at the time of enrolment in the study (estimated or known) will be used for dose calculations.

A starting dose is defined for each cohort, with dose increments for that cohort being 50% of the starting dose. The dose is reviewed and adjusted every 30 minutes as required to achieve the target heart rate (±5 beats per minute). Titration to achieve heart rate control for short lived stimulating procedures (e.g. tracheal suction, positioning, portable chest X-ray) is not required. The infusion should be continued during procedures including surgery or within hospital transfers for imaging.

Where the infusion is restarted after temporary suspension (e.g. for bradycardia or if heart rate maintained in target range without need for infusion) the starting dose for that level will be used with increments every 30 minutes as required. The infusion should not be restarted after a temporary suspension until the heart rate exceeds the minimum target rate (or 60 bpm) for more than 15 minutes.

Esmolol infusion continues according to study protocol until one of the following stopping rules is met:

- 96 hours from start of infusion
- Heart rate target achieved without esmolol for >12 hours
- Dose-limiting toxicity
- Death or withdrawal of life-sustaining treatment
- Request of participant, legal representative or treating clinician
- ICU discharge or transfer to non-participating ICU

The esmolol is weaned in steps of 5-10 mcg.kg.min^-1^ every hour from 96 hours to avoid rebound tachycardia. In the event of dose-limiting toxicity it may be reduced more quickly or stopped immediately. In the event of ICU discharge or transfer to a non-participating ICU the infusion will be reduced at a rate calculated to ensure at least two hours without esmolol infusion prior to discharge.

Bradycardia is defined for the purposes of this study as a heart rate under 50 beats per minute. The dose of esmolol should be reduced in the appropriate increments for dose level every 30 minutes until the bradycardia resolves.

Bradycardia with haemodynamic compromise is a heart rate under 50 beats per minute and a systolic blood pressure under 110 mmHg (or 100 mmHg for those aged 50-69 years). The dose of esmolol should be reduced by twice the appropriate increment for dose level every 30 minutes until bradycardia with haemodynamic compromise resolves.

Where bradycardia is severe (defined as heart rate under 30 beats per minute) the esmolol infusion may be stopped temporarily until bradycardia resolves. Further intervention is at the discretion of the clinical team including intravenous antimuscarinic or chronotropic drugs and external or transvenous pacing.

Second degree heart block without bradycardia or haemodynamic compromise should be managed by weaning esmolol as at the end of the intervention period. If haemodynamic compromise occurs, or in the event of third degree heart block, management is as for severe bradycardia.

Hypotension should be managed according to usual clinical practice taking into account cardiac status and cerebral perfusion pressure (CPP) target. Fluid resuscitation and use of vasoactive agents including catecholamine and non-catecholamine vasopressors and inodilators are permitted. Esmolol infusion should be reduced in increments if these measures are insufficient to maintain CPP.

Flow charts for the management of esmolol infusion are shown in Figures 2 and 3.

**Figure 2:**
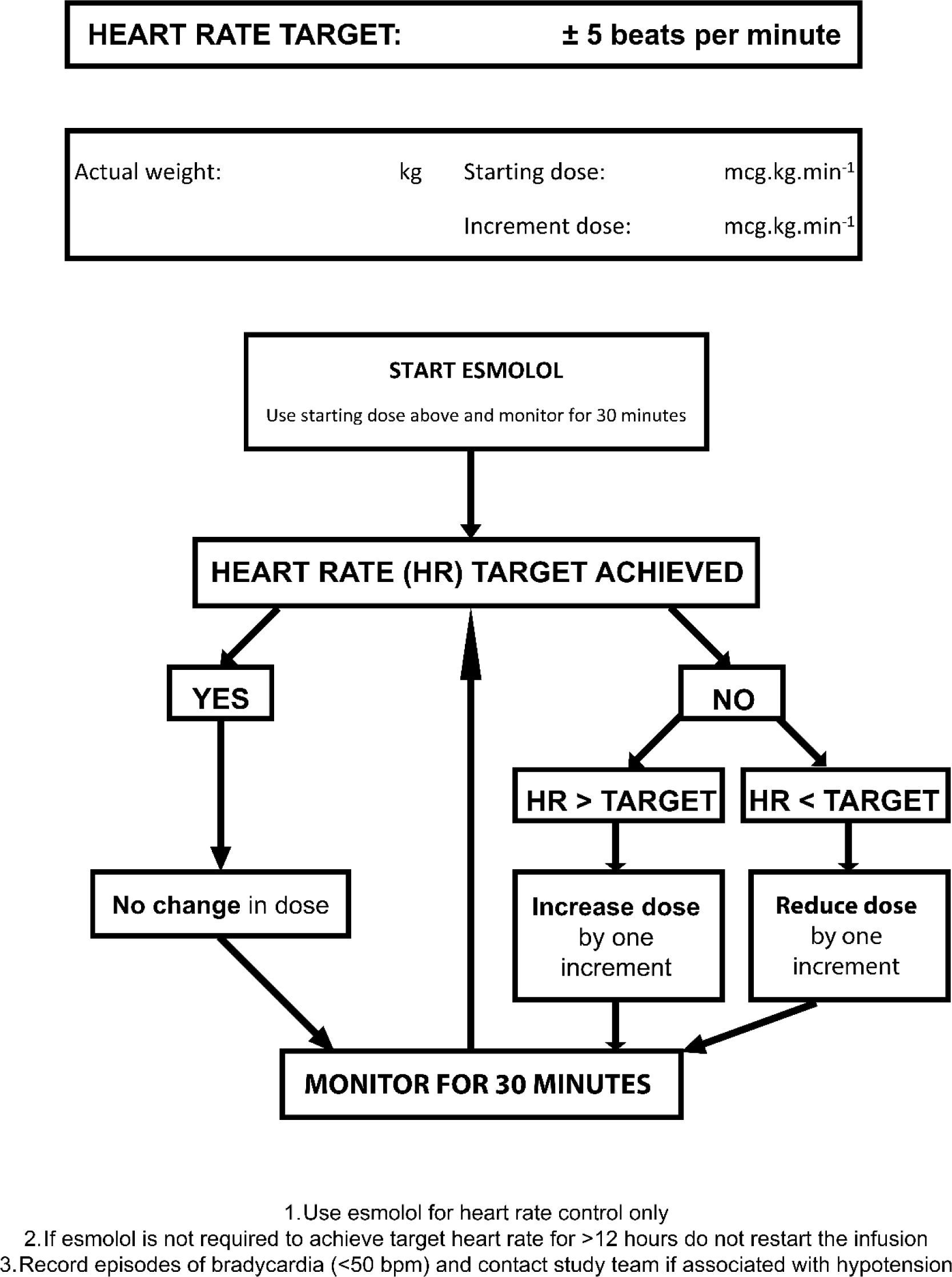
Flowchart for titration of esmolol dose. bpm – beats per minute; HR – heart rate; mcg.kg.min^-1^ – micrograms per kilogram per minute.

**Figure 3:**
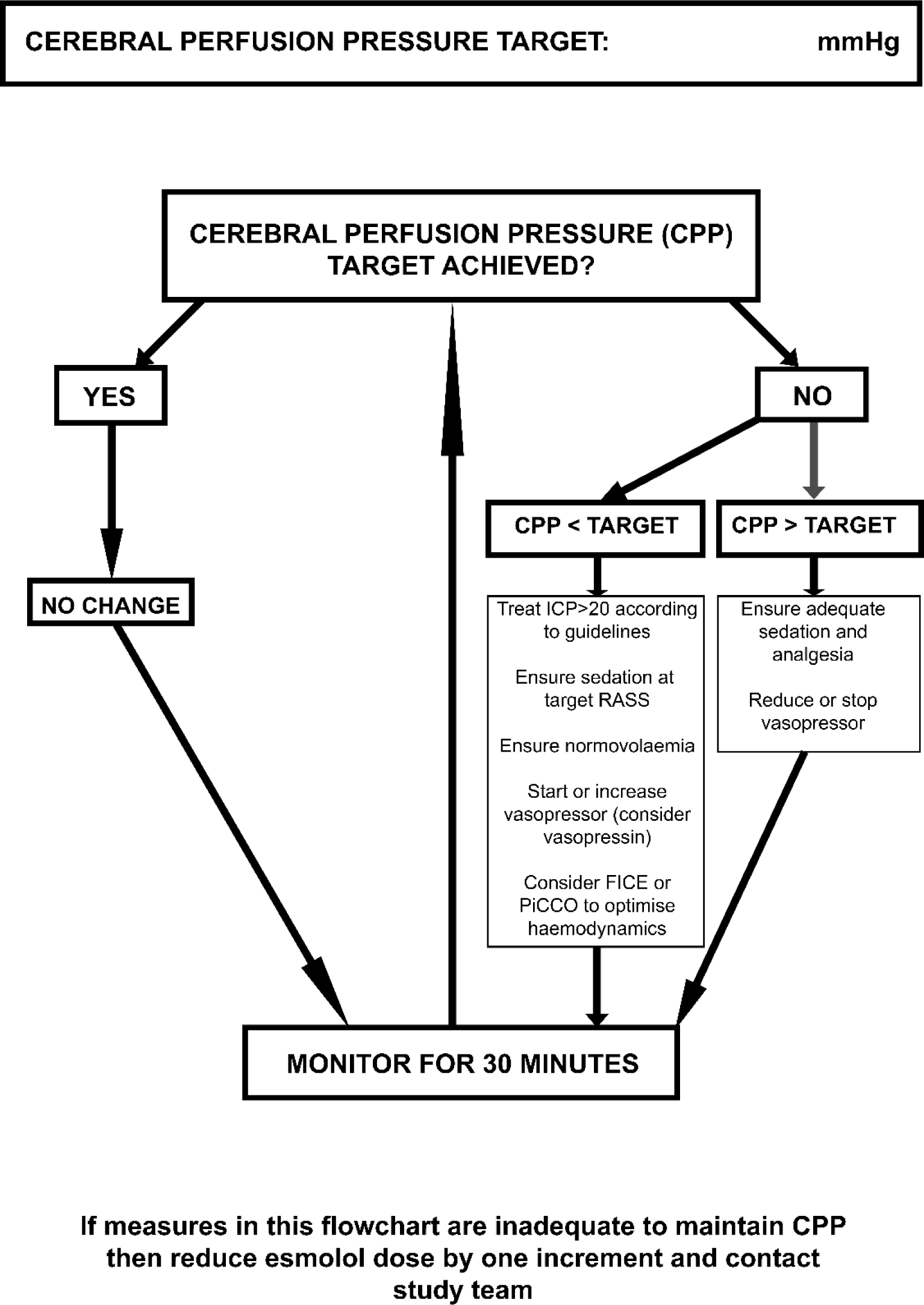
flowchart for management of cerebral perfusion pressure. CPP = cerebral perfusion pressure; FICE – focused intensive care echocardiography; ICP – intracranial pressure; mmHg – millimetres of mercury; PiCCO – pulse index continuous cardiac output; RASS – Richmond Agitation Sedation Scale.

### Concomitant interventions

Enteral or parenteral use of selective or non-selective beta-adrenergic blockers is not permitted during the intervention phase (i.e., during esmolol infusion including weaning period). Given the short elimination half -life of esmolol a two-hour gap from termination of infusion is considered sufficient. No other specifications on the use of concomitant interventions are made.

Standard traumatic brain injury management at Southmead Hospital is based on Brain Trauma Foundation Guidelines [19].

### Baseline, intervention and follow-up data

Baseline demographics collected will include age, gender, Glasgow Coma Score, time of injury and admission, Charlson comorbidity index, beta-blocker use at admission, intracranial and extracranial injury (abbreviated injury score and injury severity score) and the Helsinki CT score. Intracranial pressure directed interventions include osmotic therapy, sedation and neuromuscular blockade, hyperventilation, therapeutic hypothermia (deliberate reduction in core temperature below 35°C), radiological investigation of elevated ICP, CSF drainage and craniotomy or craniectomy. The schedule of assessments is shown in full in Table S1 (Supplementary Material).

### Primary outcome

The primary endpoint is a continual reassessment method-derived maximum tolerated dose escalation schedule for esmolol that combines clinically significant reduction in heart rate (defined as ≥15% from baseline) with maintenance of cerebral perfusion pressure.

### Secondary outcomes

Secondary outcomes in this study are:

1. Organ function
  - Sequential organ failure assessment (excluding neurological assessment)
2. Clinical
  - Mortality: ICU, acute hospital and 6-month
  - Length of stay: ICU and acute hospital
  - Duration of mechanical ventilation
  - Bloodstream infection in ICU
  - Extended Glasgow Outcome Score at 6 months
  - Quality of life (EQ-5D-5L) at 6 months

### Exploratory outcomes

Biomarkers will include cardiac troponin T (cTnT), coagulation screen, glucose, lactate and heart rate. Blood will be stored for subsequent analysis of further endothelial and other biomarkers.

Estimates of efficacy and feasibility, other than clinical outcomes listed above, will include the following:

1. Safety
  - Incidence of bradycardia (heart rate <50 beats per minute) with or without haemodynamic compromise requiring intervention other than reduction of esmolol dose
  - Incidence of second or third degree heart block with or without haemodynamic compromise requiring intervention other than reduction of esmolol dose
  - Incidence of clinically significant hypotension (systolic blood pressure <100 mmHg for patients aged 50-69 years, <110 mmHg for others) requiring intervention other than reduction of esmolol dose
2. Efficacy
  - Dose and duration of vasopressor during esmolol infusion
  - Proportion of time during esmolol infusion with cerebral perfusion pressure in target range (60-70 mmHg)
  - Number of interventions per calendar day for intracranial pressure control during esmolol infusion (with daily and domain Therapy Intensity Level scores)
3. Feasibility
  - Rates of recruitment, consent after emergency waiver and loss to follow-up
  - Non-compliance with study protocol
4. Acceptability

Additional funding will be sought for studies to support exploratory outcomes including biomarker analysis of stored blood and qualitative investigation of study acceptability and protocol delivery.

### Pharmacovigilance

Patients admitted to Intensive Care following severe traumatic brain injury are critically ill and have a high baseline risk of complications of illness and of death. Medical occurrences that meet the definition of adverse events (AEs) and adverse reactions (ARs) may be expected features of critical illness requiring ICU care. All adverse events and adverse reactions will be considered in the context of the individual patient’s clinical condition and the natural history of severe traumatic brain injury. Those that are considered by the Chief Investigator (or medically qualified designate) to be consistent with the patient’s critical illness do not require recording or reporting, unless the Investigator considers they may relate to participation in the trial. All serious adverse events (SAEs) and serious adverse reactions (SARs) both expected and unexpected will be recorded. For the purpose of this study the Reference Safety Information is the Summary of Prescribing Characteristics for Brevibloc Premixed 10mg/mL solution for infusion (Baxter Healthcare Ltd) [21].

Secondary and exploratory outcomes include assessment of organ function and significant haemodynamic side effects of esmolol infusion as a means of capturing additional safety information in the patient population. Protocol based guidance is available for the management of specific adverse events (e.g. bradycardia with haemodynamic compromise). The primary responsibility for management of adverse events lies with the treating clinician.

### Statistical analysis

Inability to maintain cerebral perfusion pressure in the presence of esmolol was chosen as the primary definition of toxicity based on the known importance of adequate blood pressure for prevention of secondary brain injury [19]. The maintenance of adequate CPP as a key goal of therapy is also familiar to clinicians practicing in the field of neurointensive care.

Only data from esmolol treated patients will be used in statistical analysis. Esmolol treated patients are those who meet all inclusion criteria and no exclusion criteria, receive any dose of esmolol within 2 hours of confirmation of eligibility and 24 hours of injury and do not withdraw consent for use of data prior to use of that data in CRM analysis.

A one-parameter logistic model, initialised with skeleton parameters as per Table 2, will be used for the CRM modelling, with estimated probabilities revised as data emerges. This likelihood modelling algorithm will identify a maximum tolerated dose (MTD) escalation schedule, with a defined prior reasoned target toxicity level (TTL), or “acceptable” toxicity rate (θ), of 10% with an indifference level of 2 percentage points for decision making. The weighting afforded to the pre-trial logistic model on estimated probabilities will be revised as trial data sequentially emerges.

**Table 2:** pre-defined dose levels for esmolol infusion. mcg.kg.min^-1^ – micrograms per kilogram per minute.

| **Dose level** | **Starting dose (mcg.kg.min<sup>-1</sup>)** | **Increment (mcg.kg.min<sup>-1</sup>)** | **Estimated prior probability of dose limiting toxicity** |
| --- | --- | --- | --- |
| 1 | 5 | 2.5 | 0.01 |
| 2 | 10 | 5 | 0.04 |
| 3 | 16 | 8 | 0.07 |
| 4 | 25 | 12.5 | 0.10 |
| 5 | 35 | 17.5 | 0.15 |
| 6 | 46 | 23 | 0.20 |
| 7 | 62 | 31 | 0.25 |

Cohort size of 3 will be used. In the absence of dose limiting toxicity (DLT), an increase in the dose escalation schedule will be considered by the Trial Management Group after the last patient in a cohort has completed the esmolol intervention period. Dose escalation cannot be by more than one level and, as an additional safety measure, the dose will not be escalated until the second cohort has completed intervention. Dosage de-escalation will be considered after each DLT and is unrestricted. Dose level changes are subject to Sponsor approval with oversight of dose decision making the responsibility of the Steering Committee comprising independent expert and lay members.

The planned sample size of 24 esmolol treated patients was determined in a pragmatic manner based on estimated recruitment rates following a test screening period in the ICU that were compatible with the timeline required by the Funder and on the expected small information gain for additional patients over and above the target. This allows testing of up to seven dose levels (Table 2).

In analysis of secondary and exploratory outcomes continuous variables will be summarised by descriptive statistics (mean and standard deviation, minimum, median, maximum and interquartile range) and categorical data will be summarised in terms of frequency and percentage.

The Sequential Organ Failure Assessment will be reported as both daily total and by variable, excluding neurological assessment. Mortality outcomes will be analysed using Kaplan-Meier survivorship. Length of stay and duration of mechanical ventilation will be analysed using time to event Kaplan-Meier analyses. Both the number of patients with a bloodstream infection and the total number of bloodstream infections in ICU will be reported. The Extended Glasgow Outcome Score will be reported by category and dichotomised into favourable (eGOS 4-8) and unfavourable (eGOS 1-3) outcome, differentiating between patients who are independent at home or not. Quality of life (EQ-5D-5L) will be reported as the EQ-5D index and by each dimension.

Blood biomarkers (troponin, glucose, lactate and INR) will be reported for each day of the esmolol infusion. The incidence of safety outcomes is the number of events per calendar day of esmolol infusion (i.e. with at least 12 hours of infusion) and will be reported as an overall incidence and by category. The dose of vasopressor will be reported as noradrenaline equivalents in micrograms per kilogram per minute, with a correction factor of 10 used to convert from metaraminol.

Rate of recruitment reported as % of screened population enrolled and as % of eligible population enrolled by month. Non-compliance with protocol, defined as need for event report to Sponsor, will be presented as total number of events and number of patients with episodes of non-compliance.

No subgroup or adjusted analyses are planned. There will be no imputation for missing data which will be recorded as missing if queries are unable to recover the data. Some types of missing data represent study outcomes and will be reported as such.

Further quantitative analysis of relationships between outcomes or other study data will be undertaken with appropriate statistical methods. Qualitative analysis of acceptability will be undertaken using constant comparison methodology adapted from grounded theory with further details available via the Open Science Framework registration (https://osf.io/9ht4v). After publication the data will be made available to other researchers on request if approved by the Trial Management Group and Sponsor.

### Funding, sponsorship and ethical review

The study is funded by the Research for Patient Benefit (RfPB) programme of the National Institute for Health and Care Research (NIHR Award PB-PG-0418-20029). The views expressed in this article are those of the authors and not necessarily those of the NIHR or the Department of Health and Social Care. The Sponsor is North Bristol NHS Trust. The study was approved by South Central – Hampshire A Research Ethics Committee (reference 20/SC/0219).

### Recruitment

The first participant enrolment occurred on 30^th^ December 2020 with the last participant final follow up expected by 30^th^ April 2023.

### Study registration

The study is registered with the ISRCTN Registry (ISRCTN11038397).

## Discussion

There is a signal suggesting a significant benefit associated with beta-blockade after severe traumatic brain injury seen in several meta-analyses, though the ideal drug and dose schedule has not been determined. The EBB-TBI research programme aims to define and test an intervention package based on esmolol. This first study aims to determine a maximum tolerated dose of esmolol given that we are aiming to institute clinically significant beta-blockade at a time where there is a real risk of exacerbating secondary brain injury through hypotension.

Although some analyses favour the use of propranolol, a non-selective beta-antagonist, over other beta-blockers the data does not come from prospective randomised trials and it is not possible to eliminate potential confounders [48,49]. Further, propranolol is compared against all other beta-blockers including other non-selective drugs as well as those with alpha-antagonist or class III anti-arrhythmic actions (labetalol and sotalol respectively). As the authors acknowledge these studies leave the question of the ideal drug open. We believe the practical advantages of esmolol together with the potential theoretical benefits make it an ideal drug for use in the early period after severe traumatic brain injury.

Use of the continual reassessment method, an adaptive model-based design for dose-finding studies, is intended to deliver a more efficient and safer study. In particular, in a patient population where the margin for error is narrow and the potential harm permanent and profound, and where the intervention challenges long held beliefs about clinical practice, minimising the exposure of patients to potential toxicity is important. Our assumptions are based on a maximum dose with only 10% acceptable probability of toxicity, against the 33% standard used in many phase 1 trials [44,50]. We believe that this approach is reasonable in this patient population given the balance of estimates of a dose that might provide benefit against one which leads to the known harm of hypotension [20,29,33,34,35].

The strengths of this study include recruitment of patients under-represented in other studies of beta-blockade after severe TBI (those with extra-cranial injuries, previous beta-blocker use or on vasopressors), early administration of intervention and an adaptive design. The intervention itself is simple and uses a drug that has rapid offset in the event of side effect or toxicity.

There are several limitations. Practice in a single centre cannot be assumed to generalise more widely. We have used a simple surrogate of sympathetic activity and have not controlled for influences such as the adequacy of sedation or fluid resuscitation. Our sample size was determined pragmatically rather than formally estimated. The clinical benefit of esmolol cannot be determined as there is no control group and some relevant outcomes will not be collected. Similarly, we have not attempted to investigate subgroups of the heterogenous TBI population that might derive particular benefit or harm from the intervention, for example based on admission troponin [51]. We plan to address these limitations in subsequent studies in the programme focused on efficacy and effectiveness.

### Conclusion

Here we present the protocol for a dose finding study of esmolol for early beta-blockade after severe traumatic brain injury using the continual reassessment method. Both drug and study design are novel in this setting. This study will determine a dose schedule for esmolol that can be tested for benefit in adults with severe traumatic brain injury.

## Data Availability

After publication the data will be made available to other researchers on request if approved by the Trial Management Group and Sponsor.

