## Supplementary Table 1 for "Early intravenous Beta-Blockade with esmolol in adults with isolated severe Traumatic Brain Injury (EBB-TBI): protocol for a phase 2a intervention design study"

| **Procedure** | **Screening** | **Start infusion** | **Infusion** | | | | **Wean infusion (if applicable)** | **Follow-up** | | |
| --- | --- | --- | --- | --- | --- | --- | --- | --- | --- | --- |
| Timeline | Day 0 | Time 0 | Day 1 (T0+24) | Day 2 | Day 3 | Day 4 | Day 5 | ICU discharge | Hospital discharge | 6 months |
| Eligibility assessment | X |  |  |  |  |  |  |  |  |  |
| Emergency waiver of consent | X |  |  |  |  |  |  |  |  |  |
| Confirmation of assent or consent to proceed |  |  | X | X | X | X | X | X | X |  |
| Demographics |  | X |  |  |  |  |  |  |  |  |
| Medical and drug history |  | X |  |  |  |  |  |  |  |  |
| Physical examination |  | X |  |  |  |  |  |  |  |  |
| Glasgow Coma Scale |  | X | X | X | X | X | X | X |  |  |
| **Procedure** | **Screening** | **Start infusion** | **Infusion** | | | | **Wean infusion (if applicable)** | **Follow-up** | | |
| Timeline | Day 0 | Time 0 | Day 1 (T0+24) | Day 2 | Day 3 | Day 4 | Day 5 | ICU discharge | Hospital discharge | 6 months |
| ECG |  | X | X | X | X | X | X |  |  |  |
| Biobank (optional) |  | X |  | X |  | X |  |  |  |  |
| Laboratory bloods |  | X | X | X | X | X | X |  |  |  |
| Arterial blood gases |  | X | X | X | X | X | X |  |  |  |
| Microbiology |  |  | X | X | X | X | X | X |  |  |
| Heart rate (hourly) |  | X | X | X | X | X | X |  |  |  |
| Cerebral perfusion pressure (hourly) |  | X | X | X | X | X | X |  |  |  |
| ICP or CPP directed interventions |  | X | X | X | X | X | X |  |  |  |
| **Procedure** | **Screening** | **Start infusion** | **Infusion** | | | | **Wean infusion (if applicable)** | **Follow-up** | | |
| Timeline | Day 0 | Time 0 | Day 1 (T0+24) | Day 2 | Day 3 | Day 4 | Day 5 | ICU discharge | Hospital discharge | 6 months |
| Vasopressor (mean daily dose) |  | X | X | X | X | X | X |  |  |  |
| Sedation (mean daily dose) |  | X | X | X | X | X | X |  |  |  |
| Fluid balance including urine output |  | X | X | X | X | X | X |  |  |  |
| Adverse events (SAEs until ICU discharge) |  | X | X | X | X | X | X | X |  |  |
| Duration mechanical ventilation |  |  |  |  |  |  |  | X |  |  |
| Length of stay |  |  |  |  |  |  |  | X | X |  |
| Mortality |  |  |  |  |  |  |  | X | X | X |
| Extended Glasgow Outcome Scale |  |  |  |  |  |  |  |  |  | X |
| EQ-5D |  |  |  |  |  |  |  |  |  | X |
| Trial end |  |  |  |  |  |  |  |  |  | X |

Table S1: schedule of assessments. CPP – cerebral perfusion pressure; ECG – electrocardiogram; EQ-5D – EuroQoL 5 Dimension; ICP – intracranial pressure; ICU – Intensive Care Unit; SAE – serious adverse event.
